# Impact of COVID-19 Pandemic on Sleep Including HRV and Physical Activity as Mediators: A Causal ML Approach

**DOI:** 10.1101/2023.06.08.23291008

**Authors:** Elahe Khatibi, Mahyar Abbasian, Iman Azimi, Sina Labbaf, Mohammad Feli, Jessica Borelli, Nikil Dutt, Amir M. Rahmani

## Abstract

Sleep quality is crucial to both mental and physical well-being. The COVID-19 pandemic, which has notably affected the population’s health worldwide, has been shown to deteriorate people’s sleep quality. Numerous studies have been conducted to evaluate the impact of the COVID-19 pandemic on sleep efficiency, investigating their relationships using correlation-based methods. These methods merely rely on learning spurious correlation rather than the causal relations among variables. Furthermore, they fail to pinpoint potential sources of bias and mediators and envision counterfactual scenarios, leading to a poor estimation. In this paper, we develop a Causal Machine Learning method, which encompasses causal discovery and causal inference components, to extract the causal relations between the COVID-19 pandemic (treatment variable) and sleep quality (outcome) and estimate the causal treatment effect, respectively. We conducted a wearable-based health monitoring study to collect data, including sleep quality, physical activity, and Heart Rate Variability (HRV) from college students before and after the COVID-19 lockdown in March 2020. Our causal discovery component generates a causal graph and pinpoints mediators in the causal model. We incorporate the strongly contributing mediators (i.e., HRV and physical activity) into our causal inference component to estimate the robust, accurate, and explainable causal effect of the pandemic on sleep quality. Finally, we validate our estimation via three refutation analysis techniques. Our experimental results indicate that the pandemic exacerbates college students’ sleep scores by 8%. Our validation results show significant p-values confirming our estimation.

## I. Introduction

In the realm of human well-being and overall quality of life, the significance of sleep quality cannot be overstated. Sleep deprivation, on the other hand, can have far-reaching consequences, contributing to a plethora of ailments encompassing obesity, depressive disorders, hypertension, coronary disease, and diabetes, among others [1]. Given the paramount significance of sleep, extensive research endeavors have been undertaken to assess various sleep parameters, including sleep duration, sleep fragmentation, and sleep efficiency. These assessments have been conducted through two primary methods: self-report questionnaires [2] and the utilization of smart wearables (e.g., the Oura ring) [3]. Moreover, recent investigations have delved into the disruptive effects of the COVID-19 pandemic on human sleep patterns [2], [3], recognizing its profound impact on the lifestyle of individuals worldwide.

Within the existing body of literature, numerous studies have been conducted to assess the influence of COVID-19 on sleep quality, employing either statistical-based methods or correlation-based machine learning models. Conventional statistical-based methods have utilized diverse techniques to examine the relationship between COVID-19 and sleep quality. For example, these methods have made use of Spearman correlation analysis to explore the associations between COVID-19 and sleep quality [5], [6]. Additionally, a maximum likelihood approach has been employed to investigate the association between sleep quality and COVID-19 [3]. In a separate study, the Pittsburgh Sleep Quality Index (PSQI) was employed as a tool to examine the impact of COVID-19 on sleep quality [5]. Each participant’s PSQI score, ranging from 0 to 21 (where lower scores indicate better sleep quality), was calculated. Subsequently, nonparametric tests were conducted, and medians, interquartile ranges (IQR), means, and standard deviations (SD) were reported in order to analyze the relationship between COVID-19 and sleep [5].

Relying solely on correlation analysis within the context of a complex phenomenon that involves multiple causal pathways presents several significant challenges. It is crucial to disentangle the causal relationships rather than relying on spurious correlations, which may result from shortcut learning between variables. In other words, methods that primarily focus on identifying and learning correlations between variables in order to estimate the impact of COVID-19 on sleep quality often fail to provide an accurate estimation of the true underlying value. Numerous studies, such as [10], have extensively explored and discussed the limitations of these methods, emphasizing the necessity of moving beyond the identification of mere data associations to comprehend the data generating process and the causal relationships among variables. For example, both statistical-based methods and machine learning models are unable to uncover the direct and indirect causal relationships between COVID-19, sleep quality, and other mediator variables, nor can they effectively detect and control for potential sources of bias.

In this paper, our objective is to examine the causal relationship and causal effect of the COVID-19 lockdown that occurred in March 2020 on the sleep quality of college students in Southern California. We take into consideration two key factors: heart rate variability (HRV) as a proxy for objective stress (autonomic nervous system activity) and physical activity. To achieve this, we introduce a novel approach called Causal Machine Learning (CML), which comprises two main components: causal discovery and causal inference. The CML method begins by employing the causal discovery component to identify the causal relationship between COVID-19 lockdown (the treatment variable) and sleep quality (the outcome variable) through the construction of causal graphs. Subsequently, the causal inference component of CML computes the causal estimations among the pandemic, sleep quality, HRV, and physical activity. To collect the necessary data, we conducted a comprehensive 12-month study [11] involving 20 college students starting in January 2020. By implementing our CML method on this collected dataset, we are able to investigate the causal impact of the COVID-19 pandemic on sleep quality. Furthermore, we validate our findings using various refutation techniques to ensure the robustness and reliability of our results.

## II. Methods

In this section, we first outline our data collection and then describe the proposed CML method.

### A. Data Collection

We conducted a health monitoring study with the objective of remotely monitoring a group of twenty college students, of whom 65% were female. The participants in our study had a mean age of 19.8 years, with a standard deviation of 1.0. This study was conducted as a part of a larger longitudinal study [11] that aimed to develop personalized models for assessing the mental and physical health of young individuals. The participants in our study were exclusively full-time college students aged between 18 and 22 years. They were unmarried, fluent in English, and users of Android phones. The study was conducted over a duration of twelve months in the year 2020, encompassing the period of the COVID-19 pandemic. The study took place in Southern California.

As part of the study, the participants were instructed to wear an Oura ring [12] conjunction with a Samsung Galaxy Active 2 watch [13] to capture various aspects of their physical activity, sleep patterns, and psychophysiological parameters such as heart rate and HRV. The Oura smart rings were specifically utilized to collect comprehensive sleep parameters, including sleep duration, wakefulness after sleep onset, and deep sleep. The ring also reported a sleep quality score, ranging from 0 to 100, served as a metric for assessing the overall quality of sleep. This score takes into account multiple factors, such as the duration of different sleep stages (e.g., light sleep), sleep efficiency, and disturbances experienced during the sleep. In addition to sleep-related data, the Oura ring also captures information about the participants’ daily activity, including movement throughout the day and rest periods. Furthermore, we incorporated the use of the Samsung watch to capture photoplethysmogram (PPG) signals at a sampling frequency of 20Hz. In our study, we employed a PPG pipeline to process the collected PPG signals. This computational pipeline involved several key steps, including the identification of highquality signals, application of appropriate filtering techniques, detection of systolic peaks, and extraction of relevant heart rate and HRV parameters [14], [16]. In our analysis, we extracted 32 HRV parameters including AVNN, RMSSD, low frequency (LF), and high frequency (HF) [16]. To facilitate the collection of data from the wearable devices, we utilized a dynamic and multi-layered platform called ZotCare [17]. This platform served as an intermediary for securely transferring data from wearables to remote servers.

This study was conducted following the ethical principles set and confirmed by University of California Irvine’s Institutional Review Board (IRB) (Health Science#2019-5153); the ethical approval was given. The participants were provided written consent prior to their participation in the study.

### B. Causal Discovery

In this study, our primary objective is to develop and employ three distinct causal discovery algorithms aimed at uncovering the underlying causal relationships between the COVID-19 lockdown and sleep quality. By utilizing these algorithms, we intend to generate concrete causal graphs that offer valuable insights into the nature of these relationships. Before applying the causal discovery algorithms, it is crucial to normalize our data to ensure that it falls within a standardized range. This normalization process serves several purposes, including promoting data uniformity, facilitating convergence during analysis, and mitigating any potential scale-related issues that may arise. Additionally, we perform standardization to calculate the covariance matrix, which is instrumental in generating the causal graphs. To capture the temporal dynamics of sleep quality accurately, we adopt an averaging approach for the HRV features, calculating the average values per day for each individual. Moreover, leveraging the temporal information provided by the data’s timestamp, we introduce a labeling mechanism. Specifically, we assign the label “COVID-19 pandemic” to the data collected after the implementation of the lockdown measures.

The employed causal discovery algorithms are Peter-Clark (PC) [18]–[20], Greedy Equivalence Search (GES) [20], and Greedy Interventional Equivalence Search (GIES) [20]– selected based on our dataset characteristics and computational resources. PC is a constrained-based causal discovery algorithm that starts from a fully-connected graph and works by assessing the conditional independence test to identify the set of candidate direct causal relations among variables [18]–[20]. PC is prominent for both its scalability and capability to tackle large-scale datasets. GES and GIES are score-based causal discovery algorithms, which start with an empty graph and add or remove edges by computing a score, such as Bayesian Information Criterion (BIC) [20]. GES can handle both linear and non-linear causal relations and diverse variable types. GIES [20] is a subsidiary of GES–the purpose of which is to diminish the computational cost.

We identify strongly contributing mediators by analyzing the generated causal graphs from our causal discovery component and adjusting for the paths. Mediator variables mediate the effect of COVID-19 pandemic on sleep quality. Note that this process was performed iteratively by taking into account the input provided by domain experts.

### C. Causal Inference

We use multiple-mediator analysis to estimate the causal effects of the treatment variable (COVID-19 pandemic) on the outcome (sleep quality score), considering the mediator variables in the obtained causal inference component. In causal inference, there are a set of statistical techniques to perform causal estimation [18], [19]. We develop a two-stage regression model for our multiple-mediator analysis: i.e., the Average Treatment Effect (ATE) and mediator analysis. ATE is a measure used to estimate the causal impact of a treatment variable on an outcome by comparing the difference in average outcomes between treatment and control groups [18], [19].

Mediate-effect analysis [19] is an important technique in causal inference, allowing us to identify the underlying mechanisms and pathways by which variables affect one another. The mediate-effect analysis examines the impact of one variable (X) on another one (Y), which is mediated by a third intermediate variable (Z), standing between these two variables (X and Y). To estimate the total effect of X on Y, each mediator analysis entails three critical components/estimations [18]: 1) the Natural Direct Effect (NDE) of X on Y, 2) the Natural Indirect Effect (NIE) of X on Y through mediator Z, and 3) the Total Effect (TE) by using the defined structural causal models [19]. TE is indeed the same as ATE which includes mediator calculations as well.

NDE refers to the expected change in Y that arises from modifying X from x to *x*^*′*^ while maintaining all mediating variables at their values under X = x, prior to the shift from x to *x*^*′*^–NDE as defined in Equation 1.

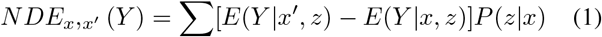

NIE of the transition from *x* to *x*^*′*^, is the expected change in Y while keeping X constant at X = x, and changing Z (for each sample row) to the value it would have reached if X had been set to X = *x*^*′*^. This counterfactual concept can be calculated as Equation 2.

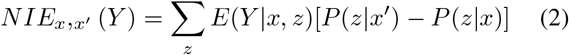

The TE of X on Y, in a Directed Acyclic Graphs, contains mediators calculated by Equation 3. In non-linear systems, the total effect is not additive. In fact, TE is the difference between the direct and indirect effects of the reverse transition (from *X* = *x*^*′*^ to *X* = *x*) [19]. In summary, TE of a transitioning from X = x to X = *x*^*′*^, where x and *x*^*′*^ can be any two levels of X (e.g., two dosage levels of a medicine), measures the expected change in the Y value caused by this change in X.

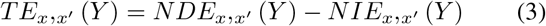

### D. Refutation Analysis

We validate the sensitivity and robustness of our estimations by examining their response to unverified assumptions by the following refutation techniques. Refutation or counterargument analysis involves introducing noise to the common cause variable or replacing the treatment with a random variable in order to challenge the estimates [18]. This analysis helps assess the reliability and stability of the estimate in the face of potential biases or confounding factors. If an estimator fails the refutation test, which means the p-value is less than 0.05, then, the estimator and assumptions are invalid. We use three refutation methods [18] in our experiments. *Placebo Treatment*: The true treatment variable is replaced with an independent random variable. The new estimate is expected to be zero; otherwise, the assumption is invalid. *Add Random Common Cause*: An independent random variable is inserted as a common cause to the dataset. The estimate is expected not to change; otherwise, the assumption is invalid. *Data Subsets*: The given dataset is substituted with a randomly selected subset. The new estimated impact is expected not to change; otherwise, the assumption is invalid.

## III Results

In this section, we present the findings of our CML methodology including the validation through the refutation analysis. Our CML method encompasses four stages for estimating causal effects: i.e., 1) constructing the causal graphs (output of causal discovery component), 2) formulating the estimand based on the causal graph, 3) employing causal estimators to compute the causal impact (outcome of causal inference component), and 4) validating the robustness of the estimate through the manipulation of assumptions.

We aim to investigate the relationship between the COVID-19 pandemic (treatment variable) and sleep quality score (outcome variable) ranging from 0 to 100. To this end, we implement our CML method on dataset (which includes 33 features) described in Section II-A. We leverage the causal discovery component to construct three causal models denoted as G_1_, G_2_, and G_3_ (see Figure 1).

**Fig. 1:**
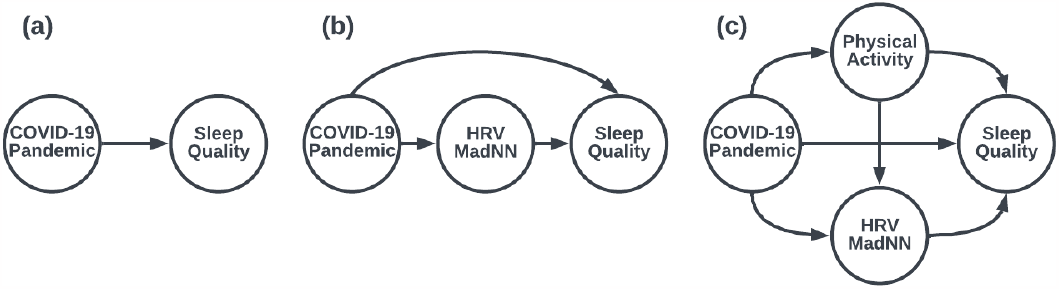
(a) Causal Graph G_1_ (b) Causal Graph G_2_ (c) Causal Graph G_3_

In G_1_ (which is our baseline model) Figure 1.a, we analyze the direct causal effect of COVID-19 on sleep which is also confirmed by causal discovery’s output. There is no confounder associated to COVID-19 pandemic. Therefore, our causal graph is confounder free. We compute the causal estimation of the treatment on the outcome, yielding a value of -0.06364. The negative sign indicates that the lockdown had a detrimental impact on the sleep quality score, Given that we standardized our data, the numerical value of -0.06364 can be interpreted as approximately 6 units of decrease within the sleep score range (0-100). To provide a meaningful interpretation of this causal estimation, we consider the sleep quality categories, reported by Oura [12], as Excellent (100-90), Good (80-89), Fair (60-79), and Poor (less than 60). This 6 unit decrease shows noticeable changes in individuals’ sleep experiences, which can change the category (e.g., from Excellent to Good). estimate for the placebo treatment was approximately zero, and the estimates for the other methods did not change, affirming the robustness of our initial estimation.

In G_2_ (Figure 1.b), we incorporate the most significant mediator –i.e., median absolute deviation of the RR interval (HRV MadNN)– extracted from the output of our causal discovery process. HRV MadNN is a measure that characterizes the variability of the interbeat interval. The inclusion of this mediator is motivated by the understanding that stress, objectively captured through HRV measurements, may contribute to the degradation of sleep quality. Consequently, we conducted a single-mediator analysis to estimate the causal effects. Our causal estimation comprises both the direct effect of the lockdown on sleep quality (NDE=-0.0781) and its indirect impact through the mediator variable (NIE=0.0160). Note that our estimation incorporates the total treatment effect (TE=-0.0941), which is computed using Equation 3.

In the causal graph G_3_ (Figure 1.c), we extend our analysis to include multiple mediators by adding the physical activity score (PA Score) as the second most significant mediator. The PA Score, reported by Oura, incorporates various factors such as steps counts, calorie burn, and intensity of activity. The inclusion of the second mediator is motivated by the understanding that the pandemic resulted in a notable increase in sedentary behavior among the general population. We applied multiple-mediators analysis which allows us to capture the combined effects of both HRV MadNN and PA Score on the causal relationship between the lockdown and sleep quality. G_3_ includes one NDE as the effect of the lockdown on the sleep quality and three NIE as 1) the effect of HRV on the sleep quality, 2) the effect of physical activity on the sleep quality, and 3) the effect of physical activity on HRV and HRV on the sleep quality. The NDE is estimated as -0.0732, and the NIE is 0.0112. Subequently, the TE is calculated as -0.0844. This result confirms that the causal impact of the pandemic on the sleep quality score is negative and significant (8.4 units). Consequently, the causal estimations of the three models highlight the adverse influence of the lockdown on sleep quality. G_1_ merely shows the direct effect of the pandemic on sleep. In contrast, G_2_ added the indirect effect of HRV, resulting in a higher TE. G_3_ also included the indirect effect of PA. However, the effect of PA on both sleep quality and HRV was removed, resulting in lower but more accurate TE.

### A. Refutation Analysis

We validate the estimated results obtained by G_3_ using the three refutation analysis methods. The obtained results are presented in Table 1. All three refutation tests validate our estimated causal effects. This is evident from the fact that all resulting P-values are greater than 0.05, indicating a lack of significant causal effects in the refutation scenarios. The new

**TABLE 1:** Validation Results for G_3_.

| Method Name | Estimated Effect | New Effect | p-value |
| --- | --- | --- | --- |
| Placebo Treatment | -0.0845 | 0.00020 | 0.61 |
| Add random cause | -0.0845 | -0.0841 | 0.47 |
| Data subsets | -0.0845 | -0.0849 | 0.43 |

## IV. Conclusion and Future Work

This paper investigated the causal effect of the COVID-19 lockdown on sleep quality among college students. Our analysis also consisted of two crucial factors, namely HRV and PA Score. To achieve this, we employed a CML method, included both causal discovery and causal inference components. We obtained insightful results that shed light on the relationship between the lockdown and students’ sleep quality. Our findings indicated that the lockdown led to an 8% deterioration in the sleep score. For future work, we will apply our CML method to assess Individualized Treatment Effects. We will also incorporate the concept of transportability into our model to facilitate the transport of causal effect to new populations.

## Data Availability

All data produced in the present study are available upon reasonable request to the authors

